# Determinants of Plaque Index Among Children Aged 5-15 Years in A Conventional vs A Religious Schools: A Cross-Sectional DAG Informed Study

**DOI:** 10.64898/2025.12.26.25343047

**Authors:** Hifza Noor, Arooba Malik, Rameesa Aymen, Mahnoor Fatima, Momina Zahra, Faisal Shafiq Malik

## Abstract

**Objective:** To compare oral hygiene status through plaque index (PI) and its determinants among children aged 5-15 years in a conventional and religious schools. This comparison was aided by Directed Acyclic Graphs (DAGs) to inform robust confounder adjustment.

**Methods:** This cross-sectional study in which 938 children were examined through the period of February 2024 to January 2025. Recorded data included demographics, sugary intake and their frequency, oral hygiene practices (dental visits, supervised toothbrushing) and clinical indicators (cavitated and non-cavitated dental lesions, proximal restoration). The outcome variable was PI taken as a continuous variable. Distribution of continuous variables was assessed through histogram and Shapiro-Wilk statistics. Bivariate comparisons utilized Mann-Whitney U (continuous non-normal) and chi-square (categorical) tests. DAGs were derived from literature and specified for minimally sufficient adjustment set criteria for multivariable analysis. Ordinary Least Square (linear regression) models were used to determine associations between outcome and predictor variables.

**Results:** In conventional school, higher PI scores were associated with older age, fewer dental visits, and absence of supervised brushing (p<0.001 for all). In religious school, PI scores were associated with age (p<0.001) and sugary intake (p=0.032). All other predictors were non significantly associated.

**Conclusions:** Determinants of plaque accumulation differ by educational settings. Supervised brushing and preventive dental visits appear important in conventional schools whereas oral hygiene practices and improved dietary assessments should be prioritized in religious schools. Lastly, DAG informed adjustments strengthen the interpretation of cross-sectional associations.

## Introduction

Early childhood caries (ECC) is a major global health challenge that affects children of all ages and ethnicities. ECC is driven by the accumulation of dental plaque which is a structured biofilm on tooth surfaces that ferments dietary carbohydrates and generates acids that demineralize enamel and dentine [1] [2].

Children aged 5 to 15 years are particularly susceptible to ECC due to their dietary habits, insufficient oral hygiene standards, and low awareness of preventive dental care [3]. Overlooking brushing habits and lack of dental checkups also contribute to ECC due to plaque accumulation. Early dental visits and oral hygiene awareness are significant in the prevention of plaque deposition, decay, gum disease, and oral infections.

Oral health status among children (of age 5=15 years) is also influenced by socio-economic perceptions and differences. Most of the families in lower- to middle-income countries (LMICs) seldom prioritize oral health screening over basic health in their routine dental visits. ECC, thus in LMICs, remains a seriously neglected health issue, attributed mainly to low prioritization and limited access and availability of dental services. A recent survey in LMICs reported high caries risk in children between 5 and 10 years of age with frequent sugary food/drink intake. It also reported that lower income families tend to present with higher plaque index (PI) scores and poor oral hygiene [4].

Early and routine dental visits are paramount in the prevention of plaque accumulation and calculus formation, all of which contribute significantly to future tooth decay, gum diseases, and oral infections [5]. Early preventive measures and improvement of good oral hygiene practices therefore greatly reducing the risks of plaque build-up.

Although literary levels are relatively high in city centres of LMICs, they still have little access to dental services [6]. A study showed that children between 6 and 12 years old in developing countries have severe oral health concerns, thus essential preventive measures need to be implemented at local level [7]. This, coupled with lack of awareness by the society about the importance of oral hygiene is a contributing factor to the oral disease burden.

Linear regression has been previously employed to investigate the association between diet, oral hygiene behaviours and socioeconomic risk factors in caries risk [8]. In order to formulate specific preventive measures to reduce the occurrence of plaque, the study determines the risk factors essential by comparing both learning institutions and enables dental experts to devise proper interventions to prevent the prevalence of plaque [9]. There are limited studies that have compared the outcome of oral health among children who attend conventional and religious schools in LMICs. This study fills the gap and brings out the high rate of plaque score in children in conventional, religious schools in urban areas of LMICs.

The hypothesis of the present study was that PI differs considerably in children according to the type of school attended because of the differences in socioeconomic background, oral hygiene behavior, and diet behavior. The aim of this research was to compare the oral hygiene status using PI and the determinants of this status among the children of 5-15 years of age in a religious and conventional schools.

## Material and Methods

This research was carried out between the period of February 2024 and January 2025. The sample size of the study was 938 children between the age of 5 and 15 years in one conventional school and one religious school including both primary and permanent dentition.

The research design was cross-sectional study involving both continuous variables and categorical variables dataset. Stratification variable was used to sort the data according to the conventional/religious schools so that it could be compared appropriately. This study design was used in order to question the variables and identify significant variations among the two categories of individuals over a specific period of time. The convenience sampling was employed, and the consent of parents was received.

Using the Raosoft calculator, a sample size of 662 was estimated [http://www.raosoft.com/samplesize.html]. The population size used in the calculation of the sample size was 200,000 with 50-percent distribution of responsibility, confidence of 99 percent, and a margin of error of 5 percent.To further reduce the margin of error to 4.2%, we took a sample size of 938 students.

### Inclusion Criteria

Children of age 5 to 15 years from both educational settings who had both primary and permanent teeth [10].

### Exclusion Criteria

Children having 10 or more missing teeth or with congenital dental anomalies (e.g. dens evaginatus, dens invaginatus, talon cusps, and supernumerary teeth) were excluded to avoid confounding results, as it influences oral health [11] [12].

The data was collected through modified and validated questionnaires from *M. Kirthiga* [13] and clinical examination. The dataset, including children from a conventional and a religious school, consists of various predictor, demographic, and outcome variables.

Demographic variables included age as a continuous variable, gender (male=0, female=1) and type of educational institution (conventional school=0, religious school=1) both as binary variables, and socio-economic status as an ordinal variable (lower=1 to upper=3) based on established scales like Kuppuswamy, BG Prasad, and Udai Pareekh’s models [20].

Predictor variables were eating habits (yes=1, no=0) and frequency of eating habits (ordinal scale=0=never, 1=occasionally, 2=frequently, 3=very frequently) which involved snack and sugary drinks. The practices of oral hygiene included dental visits (yes=1, no=0), dental practice frequency (1=none, 2=once a year, 3=twice or more a year) [15], and dental practice being supervised (yes=1, no=0) [16]. The clinical parameters of the study include cavitated/non-cavitated [17], visible plaque [18], and proximal restorations [19] and are in binary form (yes=1, no=0).

The PI is an outcome variable and a numerical value between 0 and 5 was used to represent its outcome variables. This dataset was a comprehensive examination of clinical outcomes and oral health practices which helped to find out the contributing factors.

Directed Acyclic Graphs (DAGs) were built by hand using caries-risk models where earlier research literature had identified [21], to graphically depict the relationship between PI and various contributing variables as discussed in **Figure 1**. The mediating variables such as oral health awareness and socioeconomic factors related primary exposure variables with the outcome variable.

**Figure 1:**
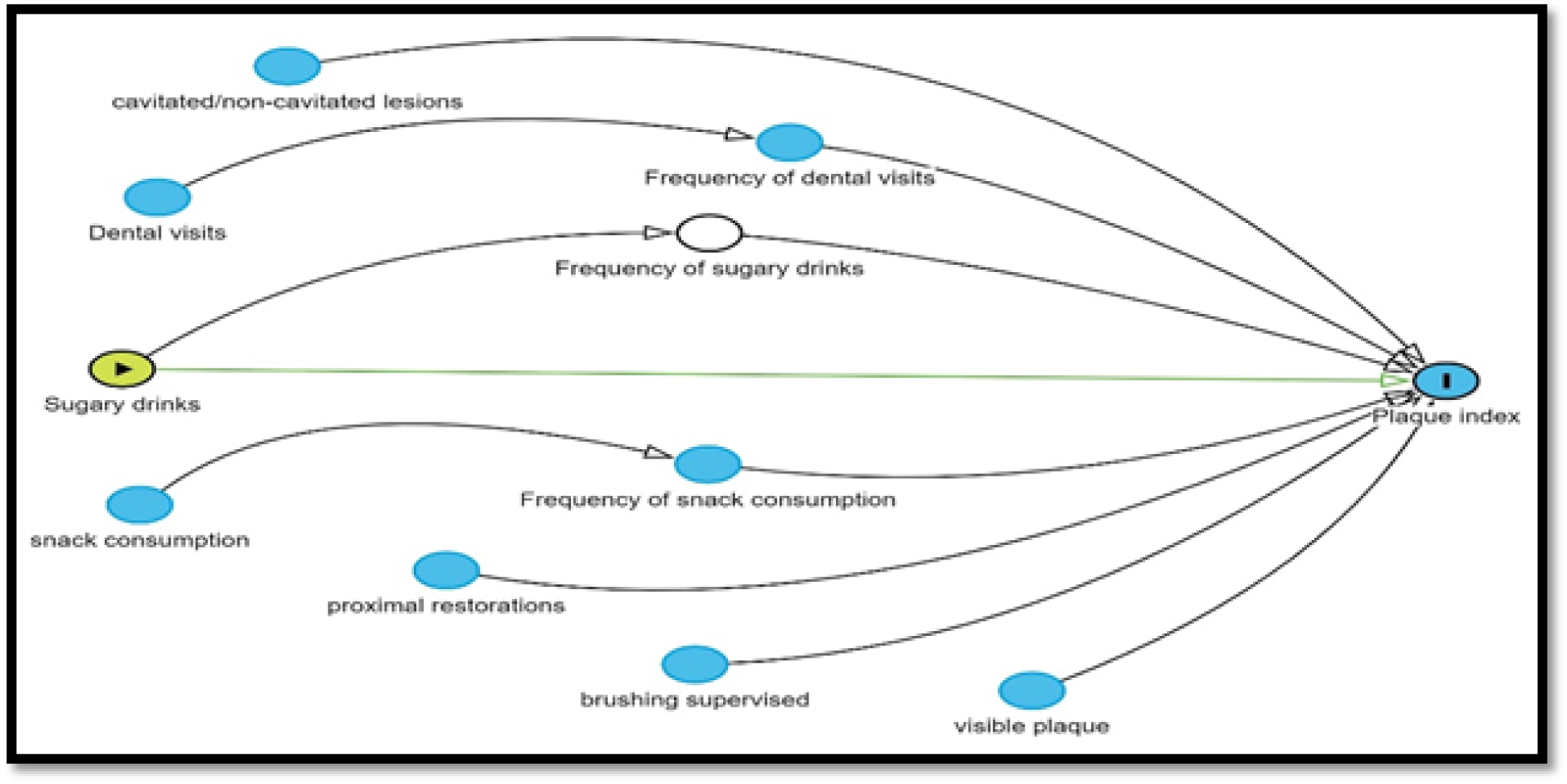
Directed Acyclic Graph (DAG) depicting the relationship between Plaque Index and various contributing variables.

In order to determine the difference between continuous and categorical variables, independent t-tests and chi-square tests were employed, respectively, and the statistical significance of p < 0.05 and the confidence intervals of 95% were used [22] and [23], respectively. The test of normality of distribution of continuous variables was conducted by the Shapiro-Wilk test. In variables that were not normally distributed, the Mann-Witney tests were applied to determine the relationship between oral health outcomes and behavior or clinical predictors in the individual school group. Multivariate linear regression models with ordinary least squares (OLS) procedures were used to find the predictors of PI in each group by using age, dietary habits, and dental visits as significant variables [24].

The visualizations utilized Python (version 3.12) libraries like *Pandas* for analyzing large datasets, *Seaborn* for statistical data visualizations and correlations, and *NumPy* for numerical calculations [25]. Statistical analyses utilized Jamovi software [26]. DAGs were made through Dagitty.net [27].

The university’s Research Ethics Committee provided ethical authorization for the study. In order to minimize selection bias, participants who matched the inclusion criteria and provided written informed consent were included in the study. On reasonable request, the corresponding author will hand over the code and dataset utilized during the research.

## Results

Univariate analysis showed most conventional school children reported high snack intake, low dental visit frequency, minimal brushing supervision, and moderate caries and plaque levels. Students in religious school (madrassa) came from lower socioeconomic backgrounds and showed higher levels of plaque and caries. **Figure 2 and Figure 3** present a univariate analysis of variables used in this study in conventional schools and religious schools (madrassa) respectively.

**Figure 2:**
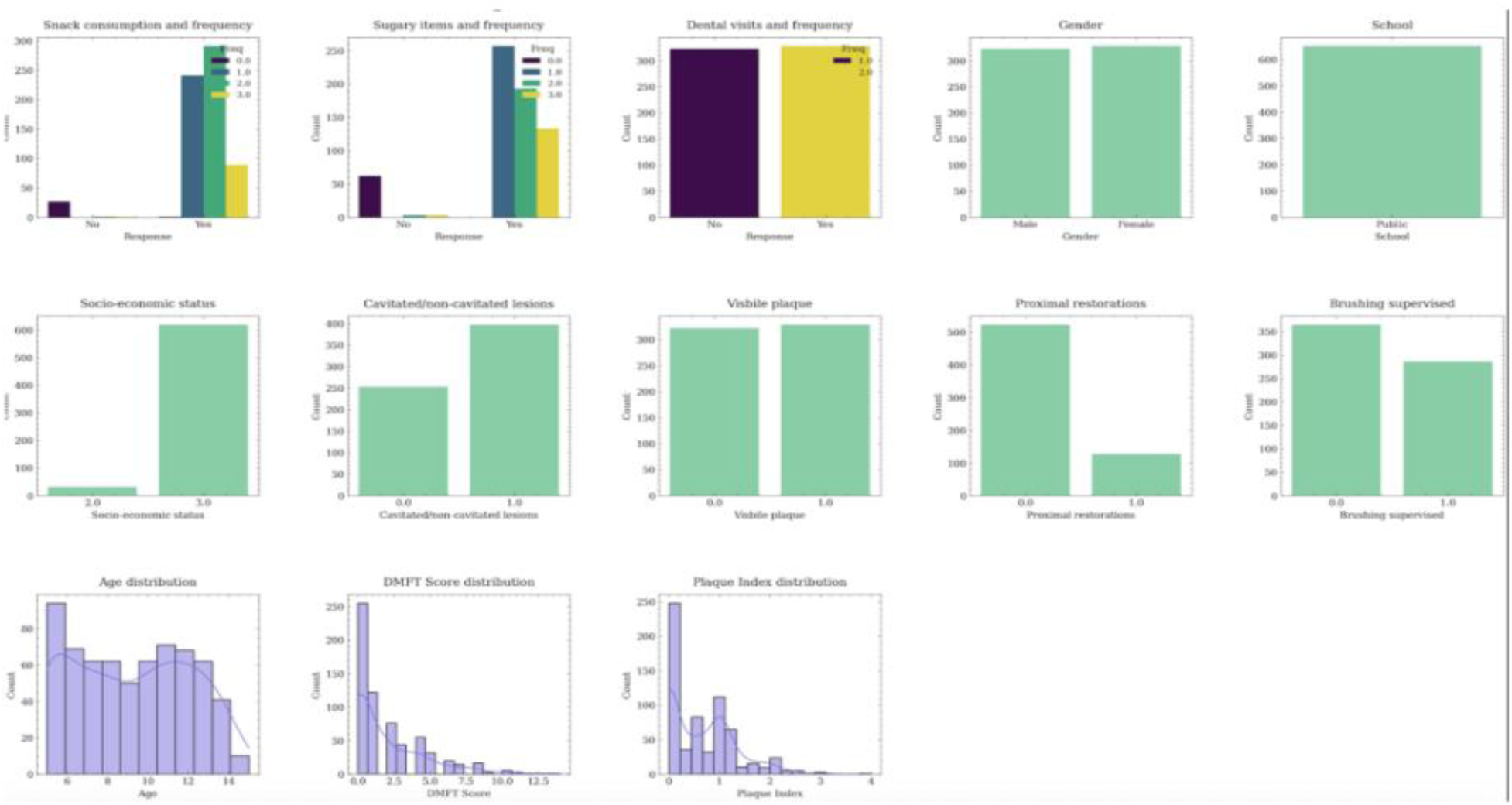
The visualization of univariate analysis of variables used in this study in conventional schools.

**Figure 3:**
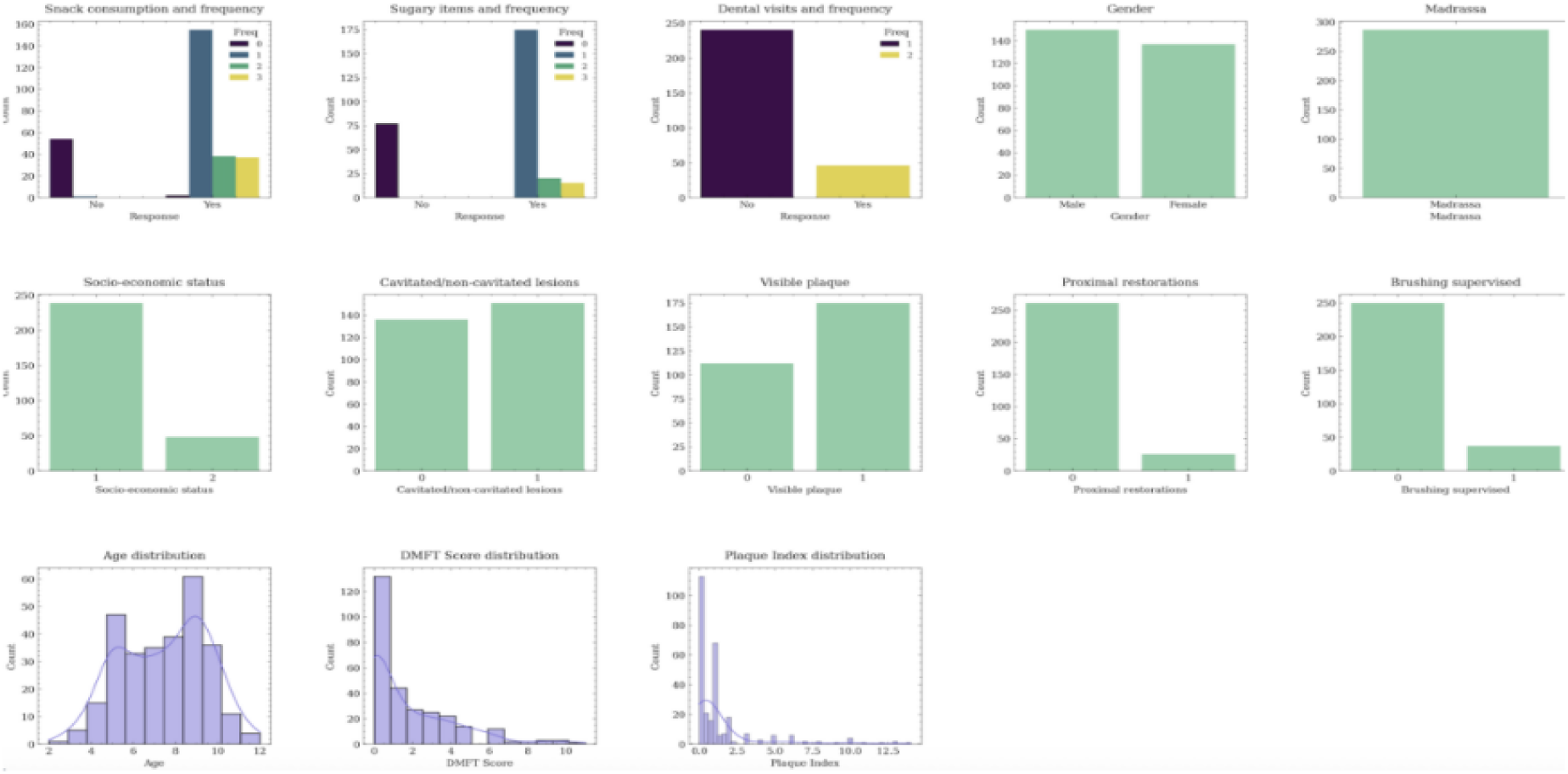
The visualization of univariable analysis of the variables of religious school (madrassa) children used in the study.

The mean age was 8.7 ± 2.85 years, with a significantly higher in conventional school (p<0.001). PI values were significantly higher among religious school students (p<0.001). Gender distribution showed no significant difference (p=0.499), indicating balanced representation across institutions. Socio-economic status varied significantly between groups (p<0.001). Dietary habits analysis indicated that religious school students reported higher frequencies of snack and sugary food consumption (p<0.001). Conventional school students were more likely to have supervised brushing (p<0.001).

The Mann-Whitney test was done because the variable of PI was not normally distributed (Shapiro-Wilk test, p<0.05). The results indicated that dental visits, cavitated/non-cavitated lesions, proximal restorations, and supervised brushing were significantly associated with PI levels (p-values=<0.001) in conventional schools (**Figure 4**). Whereas test results indicated that sugary food/drinks, cavitated/non-cavitated lesions, and proximal restorations were significantly associated with PI (p-values 0.037, 0.008, and 0.006, respectively) in religious school (**Figure 5**). Overall, these results highlight the importance of clinical and socioeconomic factors affecting oral health parameters in conventional school settings.

**Figure 4:**
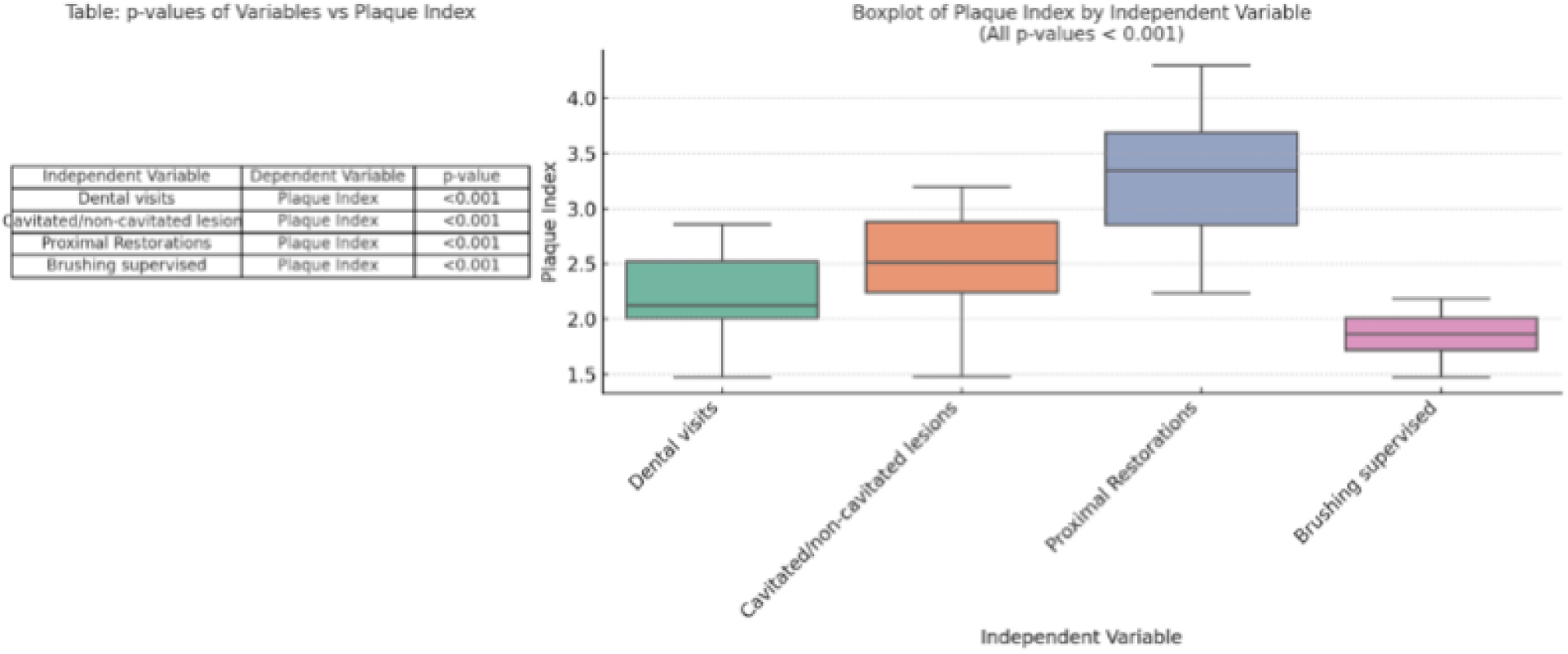
Plaque index scores of conventional school students (based on Mann Whitney test) in tabular (left) and visualization (right) form.

**Figure 5:**
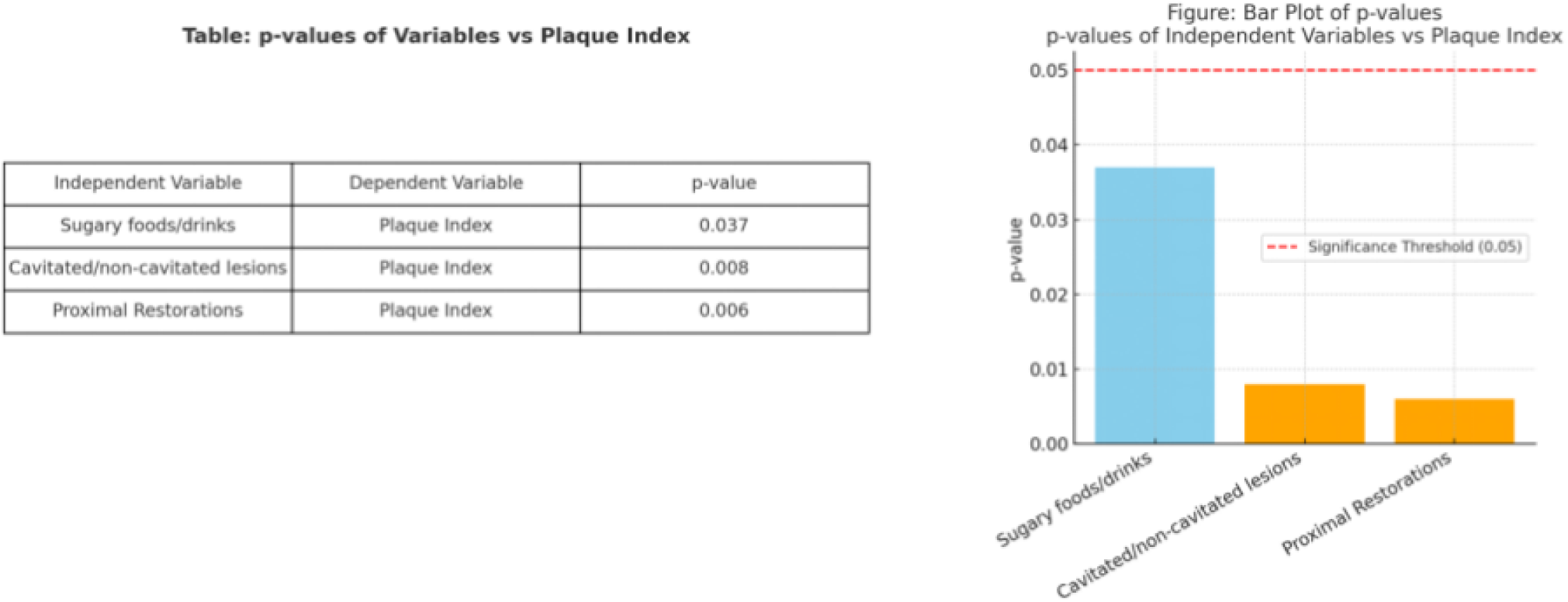
Mann-Whitney test results of religious school (madrassas) students in tabular (left) and visual (right) form.

A multiple linear regression analysis was performed to assess the influence of variables on PI. The model showed that in conventional schools, age (p<0.001), dental visits (p<0.001), and supervised brushing (p<0.001) were significant predictors. Other predictors, including snack consumption (p=0.655) and sugary food/drink intake (p=0.703), were not significantly associated with PI. **Table 1** shows linear regression results of PI as a dependent variable for conventional schools. The model revealed that in religious school, age was significantly associated with PI scores (p<0.001). Consumption of sugary foods and drinks was associated with lower PI scores (p=0.032). Other variables, including snack consumption, did not demonstrate significant associations with PI. **Table 2** shows linear regression results of PI as a dependent variable for religious school students.

**Table 1:** Linear regression results of plaque index as a dependent variable for conventional schools. * ‘No’ reference category.

| Predictor | Estimate | SE | T | P-value |
| --- | --- | --- | --- | --- |
| Intercept | 0.0683 | 0.15154 | 0.451 | 0.652 |
| Age | 0.0660 | 0.00817 | 8.071 | <0.001 |
| Snack consumption* | -0.0533 | 0.11899 | -0.448 | 0.655 |
| Sugary food/drinks* | 0.0307 | 0.08058 | 0.381 | 0.703 |
| Dental visits* | 0.1747 | 0.04874 | 3.584 | <0.001 |
| Brushing supervised* | -0.1861 | 0.04901 | -3.798 | <0.001 |

**Table 2:** Linear regression results of plaque index as dependent variable for religious school students. * ‘No’ reference category.

| Predictor | Estimate | SE | T | P-value |
| --- | --- | --- | --- | --- |
| Intercept | 0.1778 | 0.5658 | 0.314 | 0.754 |
| Age | 0.2650 | 0.0748 | 3.544 | <0.001 |
| Snack consumption* | -0.2516 | 0.4710 | -0.534 | 0.594 |
| Sugary food/drinks* | -0.9632 | 0.4460 | -2.160 | 0.032 |
| Dental visits* | 0.0456 | 0.3867 | 0.118 | 0.906 |
| Brushing supervised* | 0.7248 | 0.4317 | 1.679 | 0.094 |

## Discussion

This study evaluated PI among children from conventional and religious schools, showing that setting-specific determinants (brushing and preventive visits in conventional, hygiene and diet in religious schools) with DAG adjustments supporting casual interpretation

A significant finding of this study is that children in religious schools showed poorer oral health than those in conventional schools, likely reflecting global patterns where lower incomes, limited access to oral health education, preventive dental check-ups, or oral hygiene resources influenced oral health outcomes.[28] To close this gap, schools and communities, especially from low-income areas, need programs and policies that bring dental care and education directly to the children who need it the most.

Previous studies [6][29] consistently show that lower socioeconomic status is significantly associated with poor oral health in children. These factors may include higher rate of tooth decay, gum disease, and plaque accumulation. This can be attributed to limited dental health awareness resulting in reduced dental visits and hence reduced access to preventive care (fluoride treatments or school dental health programs).

Research has reported this significant link between age and oral health. For example, one study reported that as children progress through their school years, their oral hygiene often gets worse; they incrementally score high on plaque scores in each passing year. This pattern is especially evident when there are no school oral health programs or regular guidance from parents [30]. persistent neglect and unhealthy oral health habits seen in older age children is significantly associated with higher plaque deposition manifested in older students demonstrating higher plaque scores. This highlights the need for regular oral health awareness and dental check-ups throughout the school years. Another study reported this pattern specifically in children aged 6 to 15 years [31] [32].

The association between high plaque index and frequent dental visits, particularly in conventional schools, suggests that dental care is mostly sought reactively rather than preventively, highlighting the need for stronger oral public health promotion of regular dental checkups [32].

In these situations, going to the dentist frequently usually means the child needs treatment rather than preventive care [33]. Notably, higher dental visit frequency in such contexts often indicated underlying treatment needs, not better preventive care. Previous studies showed that children’s dental attendance was largely symptom-triggered, especially in lower socioeconomic groups, with preventive visits being rare due to lack of awareness and affordability [34].

An unexpected finding in this study was that religious school students reporting lower sugar intake had higher plaque scores likely due to reporting bias or reverse causation, misreporting of diet or high calorie foods is common among school children [35]. Reverse causation in this scenario can be implied through low sugar intake as an indicator of low socio-economic status as perceive in many LMICs, this low socioeconomic status can be linked to poor oral hygiene status as documented earlier [32].

Similarly, past studies reviewed dietary reporting in youth and found that social desirability bias and recall limitations are some of the main reasons for dietary data distortion, especially when it comes to unhealthy behaviours such as sugar snacking [36].

Reverse causality may also explain the finding, as students with prior dental problems might have reduced sugar intake giving a misleading impression of healthy oral health habits. This pattern, especially evident in low-income households, with limited dental access [37]. The limitations of self-reported underscores the need for more accurate measures such as 24-hour food recalls verified by dental investigators or tests that credible checks for dietary markers based on reliable and validated questionnaires.

## Limitations

An important limitation of this study is it being based on self-reported data. Self-reporting can be affected by participants giving socially or interviewer perceived acceptable answers. Another limitation may be that no external validation of the predictive model was performed on an external dataset.

## Conclusion

Oral health determinants such as plaque accumulation differ by educational settings and socio-economic status. Supervised tooth brushing and preventive dental visits are important in conventional schools, whereas oral hygiene practices and credible dietary assessments should be prioritized in religious schools. Lastly, causally informed adjustments based on demographic and cultural characteristics of the population should be utilized to strengthen the implementation and effectiveness of oral health programs.

## Data Availability

All data produced in the present study are available upon reasonable request to the corresponding author.

## Acknowledgement

Javed Ashraf contributed to statistical analysis.

